# Navigating Menstruation in Adolescents with Down Syndrome: A Mixed-Methods Study of Caregiver Reflections and Adolescent Voices in the UK

**DOI:** 10.64898/2026.04.22.26351080

**Authors:** Katie Greenland, Sarah Polack, Jane Wilbur

**Affiliations:** International Centre for Evidence in Disability (ICED), London School of Hygiene & Tropical Medicine, United Kingdom

**Keywords:** menstrual health, menarche, Down syndrome, adolescent, intellectual disability

## Abstract

Adolescents with Down syndrome face unique menstrual health challenges, yet their experiences remain under-researched. This study aimed to investigate the menstrual health and wellbeing of adolescents with Down syndrome in the UK, examined primarily through the reflective lens of primary caregivers alongside direct accounts from adolescents, to inform the development of tailored, evidence-based interventions for this population.

Guided by an advisory group of caregivers and young people with Down syndrome, this mixed-methods study (September 2024 – September 2025) involved a national online survey of primary caregivers (N=143) and participatory interviews with adolescents (n=6), mothers (n=11) and healthcare and education professionals (n=8). Quantitative data were analysed descriptively according to support needs (high vs low), and qualitative data were analysed thematically.

The median age of menarche (12 years) aligned with the general population. While adolescents generally coped better with menarche than caregivers anticipated, 91% of 120 caregivers of adolescents who had reached menarche had ongoing menstruation concerns. While products like period underwear (“*magic pants”*) improved independence and simplified care, key remaining concerns included: heavy periods (48%); personal care (45%); menstrual pain (45%); and the communication of pain (26%). The impact on adolescent wellbeing was greater for those with higher support needs. Additionally, 33% of caregivers felt “overwhelmed” by menstrual-related care. Decision-making about hormonal intervention was a source of heavy responsibility for caregivers. There is substantial demand for accessible educational and practical resources to support menstruation.

Menstrual health is a highly individualised experience for adolescents with Down syndrome. Significant unmet needs persist, particularly for those with higher support needs. Successful outcomes require supporting caregivers through provision of accurate information that dispels pre-menarche anxiety alongside accessible and appropriate guidance to foster young people’s independence, choice and autonomy. Future interventions must be co-developed with the Down syndrome community to ensure safe, dignified menstruation.

Funding: Down’s Syndrome Research Foundation UK

## Introduction

Societal narratives often describe periods as a “shameful secret”, rather than a normal biological function. Recent activism (1), increased focus on women’s health (2), and specific inclusion in the Sustainable Development Goals (3, 4) have begun to reduce stigma about menstruation. This has led to open discussions about challenges like “period poverty” (5), lack of access to affordable and appropriate pain medication, and the need for adequate menstrual materials and facilities, from a social justice perspective (6). Consequently, menstrual health is increasingly recognised as a holistic “state of complete physical, mental, and social well-being” (7).

Despite growing research into the impact of menstruation on the general population (8, 9), remarkably little focus has been comparatively given to the menstrual health of people with intellectual (learning) disabilities (10, 11). Intellectual disability is characterised by significant limitations both in intellectual functioning and in adaptive behaviour, originating during the development period (12). This study focuses on Down syndrome, the most common genetic cause of intellectual disability, which is frequently associated with physical comorbidities that may uniquely influence the menstrual experience (13).

Evidence indicates that people with intellectual disabilities face disparities in their menstrual health experiences (11), mirroring broader health and healthcare inequalities (14). Potential challenges include a limited understanding of the menstrual cycle and pre-menstrual syndrome (PMS) symptoms (10, 11), difficulties with self-care (15) (often complicated by mobility, dexterity, or continence issues (16)) and barriers to communicating pain and needs, with implications for wellbeing (10, 11, 17, 18). There are also reports that some groups experience heavy, irregular or lengthy periods (19–21), which can complicate care (22, 23). For some, hormonal changes are associated with increased seizures (16, 24) and changes in behaviour (25). These issues can be compounded by “diagnostic overshadowing”, with menstrual symptoms incorrectly attributed to a person’s disability (11). Within the UK context, foundational research by Mason and Cunningham (2008) identified many of these challenges among women with Down syndrome (21). Menstruation challenges can also have a substantial impact on caregiver wellbeing (10, 26).

The onset of periods can be a particularly difficult time for adolescents with an intellectual disability and their caregivers (23), yet existing research is often dated and fails to reflect the experiences of young people with intellectual disabilities today (11). Recent reviews have identified significant gaps in research and support of menstrual health for people with intellectual disabilities, and have highlighted the need for co-produced, accessible information and communication tools (10, 11). We conducted this study to explore the menstruation experiences of adolescents with Down syndrome and their caregivers in the UK to identify unmet requirements and opportunities to improve menstrual health outcomes for people with Down syndrome and the broader intellectual disability population.

## Methods

### Study design

This mixed-methods study involved a national online survey of primary caregivers of adolescents with Down syndrome, followed by in-depth interviews with adolescent girls with Down syndrome and their family and professional caregivers.

### Patient and public involvement

The study was guided by an Advisory Group comprised of medical experts, educators, young women with Down syndrome, and family caregivers. The group was established prior to protocol development, and all members were financially compensated for their expertise and time. Contributions were gathered iteratively through online meetings and email feedback, with the highest intensity of involvement occurring during the study’s preparatory and design phases. To facilitate meaningful participation, young people with Down syndrome were supported by their caregivers (their mothers), though one woman with Down syndrome participated in meetings independently. Accommodations to maximise accessibility included circulating simplified information prior to meetings, allowing ample time for responses, offering a “come-and-go” meeting structure, and having caregivers help rephrase concepts in real-time. Participants were encouraged to have their camera on for online meetings if they felt comfortable to do so.

The Advisory Group co-designed and shaped several core components of the study methodology. Members identified priority research topics, shaped recruitment strategies for the online survey, and advised on acceptable locations for adolescent interviews. The group also reviewed and piloted the Easy Read information sheets, capacity-to-consent checklists, and assent forms, resulting in substantial modifications to eliminate ambiguous or complicated language. At the explicit suggestion of the Advisory Group to assist participants unfamiliar with research, a short video was produced to explain the study. The video demonstrated the researcher entering a home and conducting an interview alongside a trusted adult. The group also reviewed and piloted the online survey, interview guides, and visual aids used in interviews with adolescents, improving the clarity and relevance of both the questions and the response categories. The Advisory Group also contributed to discussions on subgroup analysis and reviewed the final Easy Read study outputs and dissemination formats to ensure findings were accessible to young people with Down syndrome.

#### Study population and recruitment

The online survey targeted primary caregivers of girls with Down syndrome aged 10-19 years in the UK, with recruitment from 18/09/2024 to 23/07/2025. This was an exploratory study aiming to describe menstruation experiences in a historically under-research population, so a formal sample size calculation was not performed. To maximise reach and sample diversity in terms of location and ethnicity, recruitment occurred via social media (Facebook) and the websites of four national (Down Syndrome UK, Down Syndrome Scotland, the Down’s Syndrome Association, and the Down’s Syndrome Research Foundation) and 90 local Down syndrome organisations across the UK, with recruitment drives at regular intervals during the data collection period.

For the qualitative component (in-depth interviews), we aimed to sample up to 10 adolescents, 10–15 family caregivers, and up to 15 professionals. These targets were determined by balancing anticipated data saturation against available institutional resources. Adolescents aged 13-19 with Down syndrome who were at least three months post-menarche were identified through local Down syndrome groups in London and surrounding counties to enable in-person interviews to take place. While we aimed for a sample reflecting a range of ages, levels of independence and communication skills, all individuals who met the inclusion criteria and provided informed consent within the study timeline were enrolled. No exclusions were made *a priori*, unless caregivers indicated that an interview would not be feasible for the adolescent. Primary caregivers of participating adolescents were also invited to participate. A proxy interview was conducted with the caregiver when it was not possible to interview an adolescent directly, ensuring the perspectives of families with more complex disabilities were included. Medical and education sector professionals were predominantly identified through caregiver referrals; practitioners who shared their contact details were approached for interview. In-depth interview recruitment occurred between 24/02/2025 and 09/09/2025.

### Data collection procedures

Study tools were structured around the socio-ecological framework for Menstrual Hygiene Management (MHM), adapted for disability (27), and the integrated model of menstrual experience (9). These frameworks ensured a comprehensive investigation across personal, interpersonal, societal, environmental, and biological domains. Specific questions (in the survey, and qualitative interview guides) aligned with the framework domains were informed by the literature and refined following piloting by the Advisory Group (Table S1).

The online survey was developed using Open Data Kit (ODK) and was available in English and Welsh. Data were captured on socio-demographics, preparedness, caregiver concerns and future information needs. For participants whose daughters had reached menarche, additional questions were asked on menstruation experiences and adolescent and caregiver wellbeing.

Adolescent in-depth interviews were conducted face-to-face in their homes using participatory methods tailored to individual cognitive and communication needs. Visual aids included physical period products (e.g. disposable pads and period underwear), images depicting a range of positive and negative emotions to help participants express how they feel during menstruation, and Augmentative and Alternative Communication (AAC) devices were used by participants when necessary. In addition, interviews used the “Bishesta doll” developed by Wilbur et al. (28) for menstrual health research with individuals with intellectual disabilities in Nepal. The doll has removable clothes, underwear and a detachable absorbent pad (used and unused), and accessories such as a hot water bottle. Engagement with the visual aids was driven by individual preferences. For some participants, the doll served primarily as a source of comfort during the interview. For others, the doll and its accessories helped prompt description of menstruation routines, aspects of independent management, and a prompt for discussion about pain and pain management. Adolescents’ reactions to the used menstrual absorbents enabled the research team to explore attitudes to periods and menstrual blood. Some participants did not engage with the doll as they thought the doll was for younger children. All adolescent interviews took place in the presence of a trusted adult (in all cases, the participant’s mother). Researchers (KG, SP, JW) conducted interviews in pairs whenever possible. All researchers are experienced in communicating with individuals with intellectual disabilities; KG and JW are trained in Makaton sign language and JW has worked with AAC devices. Interviews with adolescents lasted from eight to 25 minutes, concluding either when the participant indicated the interview was over or when the discussion had concluded.

In-depth interviews with primary caregivers and professionals were conducted face-to-face or via Zoom. When both were interviewed, caregiver interviews followed adolescent interviews to provide context and allow for clarifications. Adolescent interviews focused on emotions, pain, navigating menstruation, and home and educational support. Caregiver and professional interviews also covered healthcare interactions, hormonal medication, and family wellbeing. All interviews were audio-recorded when consent was provided, or documented via detailed notes when it was not.

### Research positionality

In-depth interviews were led by the authors, who are experienced in menstrual health and disability research, including with adolescents with Down syndrome. As one author is also the parent of a child with Down syndrome, we mitigated potential bias through regular team discussions to critically reflect on emerging findings and refine lines of inquiry.

### Data analysis

Quantitative survey data were analysed in *Stata 18* using frequencies and proportions. Data from caregivers of adolescents who had reached menarche and those who had not were analysed separately. Descriptive analyses for the post-menarche population were stratified by level of support needs, with ‘Lower support needs’ defined as complete independence, or minimal to moderate support required for personal care tasks (e.g., toileting, showering and dressing) and ‘higher support needs’ defined as requiring significant or full assistance with personal care, and/or formal dual diagnosis of Autism Spectrum Disorder (ASD). When co-occurring with Down syndrome, dual diagnosis with ASD typically results in sensory processing and verbal communication differences associated with higher support needs (29). Comparisons by support needs were performed using Chi-squared tests or Fisher’s exact tests.

Qualitative data in the form of interview notes and *verbatim* transcripts were checked for accuracy, deidentified by the research team, and managed using *Nvivo 15* software. Data were analysed thematically using both deductive and inductive coding techniques: Researchers (KG, JW) immersed themselves in the data by reading all transcripts and notes multiple times prior to formal coding. The research team developed a codebook informed by the study’s conceptual frameworks (Table S2) and used open-ended coding to capture emerging themes and outliers. Text segments were cross-coded to multiple codes as necessary. Coding was performed collaboratively within a secure, shared *NVivo* project file. Progress was shared iteratively among the co-authors through regular meetings to review each other’s coding and discuss the data. Once initial coding was complete, the researchers met in person to synthesise and summarise the data within each thematic node, using a thematic framework structured around the codebook. Within this framework, the team ensured data were triangulated, documented recurring patterns, anomalies, unexpected findings and initial interpretations. The research team subsequently reviewed these mapped summaries to refine overarching themes and construct the narrative for the results section. Pertinent quotes were identified for inclusion in outputs, with pseudonyms used to protect participant anonymity.

Findings were synthesised and are presented together within thematic headings, with qualitative narratives used to elaborate on and contextualise statistical survey trends. To translate these integrated findings into actionable outcomes, the research team mapped the identified unmet needs, systemic barriers, and practical successes across both datasets to construct four strategic pillars for policy and practice, which are detailed under the Implications section in the Discussion.

### Ethics and consent procedures

Ethical approval was granted by London School of Hygiene & Tropical Medicine (Ref: 31109) and the Health Research Authority (HRA) Social Care Research Ethics Committee (London - Brighton & Sussex) (Ref:24/LO/0493).

Survey participants provided digital consent before accessing the survey. Written informed consent was obtained from all interviewed caregivers and professionals. A short film explaining the study was circulated to adolescent participants a week prior to interview alongside an Easy Read information sheet. Researchers formally assessed capacity to provide informed consent for participants aged 16–19 following a face-to-face review of this Easy Read material. This assessment involved a four-item comprehension check developed in collaboration with the study’s Advisory Group. To establish capacity, participants were required to demonstrate a basic understanding of the topic (periods), study expectations (answering questions), the voluntary nature of participation (they do not have to do the interview), and their right to stop the interview at any point. Formal written informed consent was obtained if the participant demonstrated understanding of all four capacity areas. Assent was sought when a participant was deemed to lack capacity to consent, in which case the primary caregiver signed a consultee declaration form. Written parental consent and adolescent assent were obtained for participants under the age of 16. To ensure assent was informed, understanding was verified using a simplified subset of the comprehension questions (*“What will we talk about?”* and *“Do you have to do it?”*). If a participant was unable to satisfy the assent criteria, or if they actively dissented or showed any ambiguity, the adolescent interview did not proceed and a proxy interview with the parent took place instead.

## Results

Quantitative and qualitative results are integrated and presented thematically across three themes: 1) preparation for menarche; 2) menstrual experiences; and 3) support systems and information demand.

### Participant Characteristics

In total, 143 caregivers of adolescents aged 10–19 years with Down syndrome completed the survey within the recruitment window. The majority of the adolescents represented (84%, n=120) were menstruating (median age 12 years: interquartile range 11-13; range 8-16), with 68% (n=81) being at least two years post-menarche. Respondents were predominantly female (99%, n=142). The sample was primarily White (86%, n=123), but provided broad geographical representation across the UK. The sample represents diverse educational settings and functional abilities; 28% (n=40) had high support needs (Table 1).

**Table 1.** Socio-demographic and functional characteristics of the young people with Down syndrome in the caregiver survey (N=143)

|  | n (%) |
| --- | --- |
| **Caregiver relationship to child** |  |
| Female Parent / Legal Guardian | 142 (99.3%) |
| Male Parent / Legal Guardian | 1 (0.7%) |
| **Child's age (years), median (IQR) [range]** | 15 (12 – 17) [10 – 19] |
| **Ethnic group** |  |
| White | 123 (86.0%) |
| Mixed | 9 (6.3%) |
| Asian or British Asian | 4 (2.8%) |
| Black or Black British | 0 |
| Arab | 1 (0.7%) |
| Prefer not to say | 1 (0.7%) |
| **Region** |  |
| England – North | 40 (28.0%) |
| England -Midlands | 17 (11.9%) |
| England – South | 48 (33.6%) |
| Wales | 6 (4.2%) |
| Scotland | 13 (9.1%) |
| Northern Ireland | 0 |
| Prefer not to say | 1 (0.7%) |
| **Educational setting** |  |
| Mainstream school or college | 64 (44.8%) |
| Mainstream school or college with special unit / facilities | 19 (13.3%) |
| Special school, specialist college or vocational centre | 68 (47.6%) |
| Other | 9 (6.3%) |
| **Has started menstruating** |  |
| Yes | 120 (83.9%) |
| No | 23 (16.1%) |
| **Ability to express needs and wants** |  |
| Consistently uses words | 99 (69.2%) |
| Uses a combination of words, signs and gestures | 36 (25.2%) |
| Relies on gestures/has difficulty expressing needs | 8 (5.6%) |
| **Independence with personal care tasks\*** |  |
| High independence (Independent/minimal assistance) | 58 (40.6%) |
| Requires moderate assistance | 54 (37.8%) |
| High support needs (significant/full assistance) | 22 (15.4%) |

| **Formal diagnosis of Autism Spectrum Disorder (ASD)** |  |
| --- | --- |
| Yes | 19 (13.3%) |
| No | 124 (86.7%) |
| **Support needs** |  |
| Low (requires no, minimal or moderate assistance with personal care tasks) | 103 (72.0%) |
| High (significant or full assistance with personal care tasks) and/or formal diagnosis of ASD | 40 (28.0%) |

IQR = interquartile range. Other educational setting: home schooled (n=3); EOTAS or awaiting EOTAS (n=3); not in education (n=3).
\* Personal care defined as tasks including toileting, brushing teeth, showering and dressing.
ASD = Autism Spectrum Disorder

Twenty-five in-depth interviews were conducted across three participant types. First, interviews were completed with six adolescents with Down syndrome aged 13–19 (median 17.5 years), including two individuals with a dual diagnosis of Down syndrome and ASD. An additional three adolescents declined to participate either during the consenting process or at the start of the interview; all three individuals had limited verbal communication skills and signalled their disinclination or discomfort by choosing not to remain in the interview room, in which case interviews did not take place. Second, interviews were conducted with 11 mothers, namely the primary caregivers of the six participating adolescents, the three adolescents who declined to be interviewed, and two who provided proxy interviews for individuals with complex needs, totalling five caregivers of adolescents with a dual diagnosis of ASD). Finally, eight professional interviews were conducted, comprising a school nurse, a general practitioner (GP), a paediatrician, and five educators from mainstream and special schools. The profile of adolescents interviewed directly or described by their caregivers is shown in Table 2.

**Table 2.** Characteristics of Caregiver and Adolescent Interview Participants (N=17)

| Characteristic | Subgroup | Count (n) |
| --- | --- | --- |
| Participant Type* | Caregiver & Adolescent | 12 (6 dyads) |
|  | Caregiver only | 5 |
| Adolescent Age | 13-15 years | 3 |
|  | 16-19 years | 8 |
| Age at Menarche | Median (range) | 13 years ( 9-15 years) |
| Current Educational Setting | Mainstream school | 2 |
|  | Special school | 8 |
|  | Specialist unit in mainstream college | 1 |
| Diagnosis | Down syndrome only | 6 |
|  | Down syndrome and Autism Spectrum Disorder (ASD) | 5 |

\* When adolescents and their caregivers were both interviewed, the caregiver interview followed the adolescent interview, even though caregivers were physically present during adolescent interviews.

Quantitative analysis in the following sections focusses on the 120 caregivers whose daughters were post-menarche at the time of the survey, unless otherwise stated.

### Caregiver concerns and preparation for menarche

Caregivers anticipated menarche with concern: 62% (74/120) of survey respondents were moderately or very worried about how they and their child would manage menstruation. Severe worry was significantly higher among caregivers of adolescents with higher support needs (51% very worried vs. 28% lower support needs; *p*=0.015). Primary concerns were navigating menstruation (using products and personal care) (88%, n=106) and the adolescent’s response to blood (80%, n=96) (Figure 1). Caregivers of children with higher support needs were also disproportionately concerned about dealing with menstrual blood (80% vs 59%, *p*=0.028), communicating pain (69% vs 40%, *p*=0.004), social appropriateness (67% vs 47%, *p*=0.032), and hormonal impacts on mood and behaviour (66% vs 42%, *p*=0.020).

**Figure 1.**
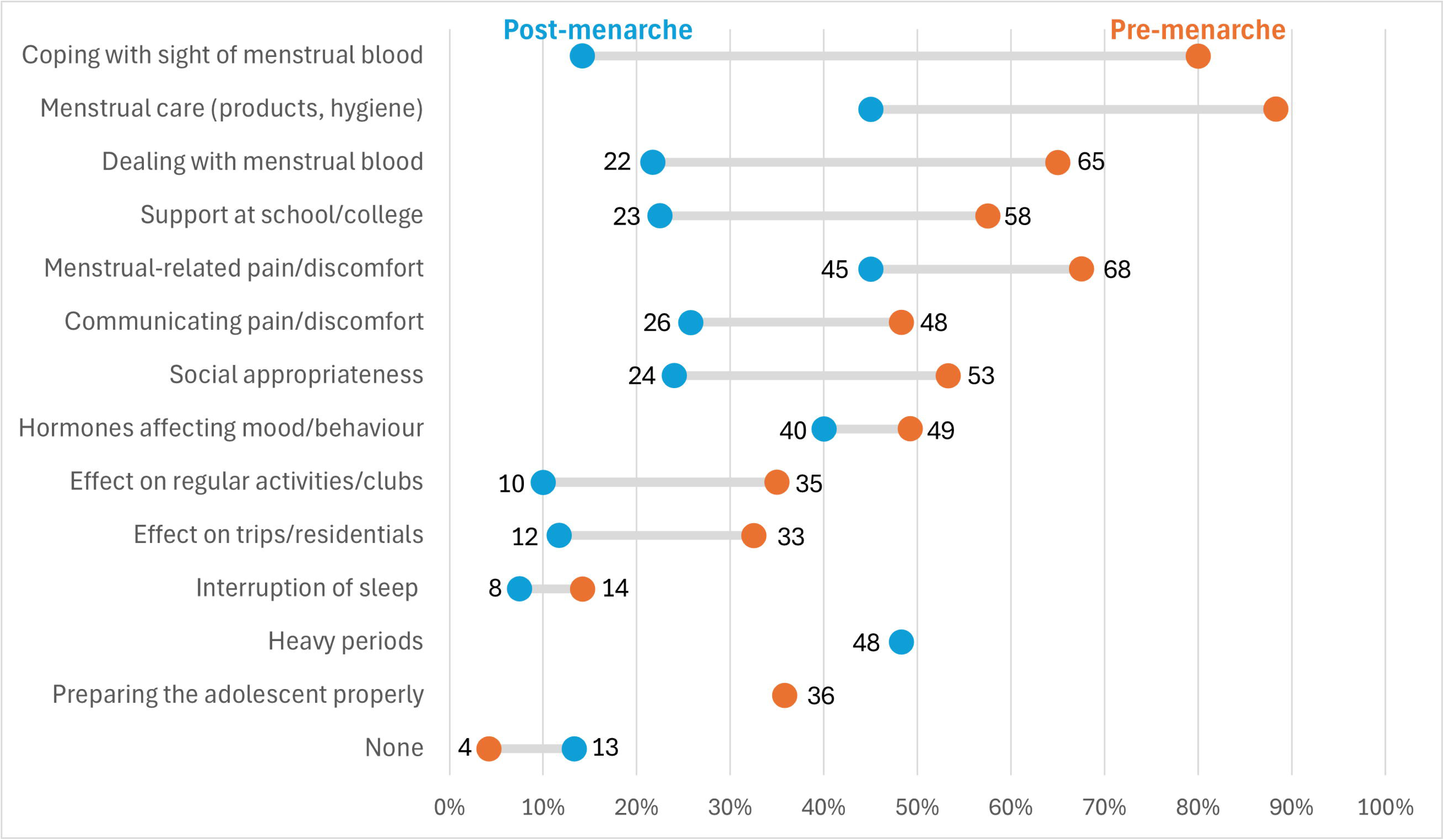
Pre- and post-menarche concerns of caregivers (N=120) Footnote: Caregivers were only asked about concerns regarding ‘preparing them properly’ pre-menarche, while ‘heavy periods’ was exclusively asked about post-menarche.

Although 59% (n=71) of survey respondents discussed menstruation at home prior to menarche to normalise periods, many caregivers explained during interview that advance conversation would have had limited use. As Emma remarked: *“It’s impossible to explain something she hasn’t experienced to her.”* Preparation focused on experiential learning and “open-door” policies: *“I’ve always been open with me and my body… bathroom doors are open”* (Joanne). Early introduction of menstrual products helped build familiarity and tolerance, as explained by Diane:

> *“I bought those really thin panty liners as a starter. So I was aware that she will be starting her period soon, so I tried to get her used to wearing like sanitary towels… it’s like wearing nothing, but they’re there, and then every time she goes to the toilet she’ll see it …”*

We observed uncertainty and some misconceptions regarding the timing of menarche. Nicole reported considerable distress after a doctor suggested early menarche (age 10) would stunt her daughter’s growth:

> *“[The doctor] said… ‘when girls get their period, they pretty much stop growing… and people with Down syndrome don’t grow tall generally.’ [Name] started her period, and she was literally still quite diddy … that was one of the most upsetting things about it”*.

Despite initial anxiety, most study participants reported that their child handled menarche better than they had expected. Overall, 58% (n=70) of survey participants reported that their child took the onset of menstruation in their stride or reacted positively, while 18% were worried, 12% were surprised/confused, and 11% were scared or distressed. The prevalence of negative emotional reactions was similar regardless of whether prior discussions about menstruation had taken place. Nicole reflected that she had feared her daughter would *“freak out”*, or *“think that she’s going to die”,* yet she *“just seems to take it in her stride.”* Nicole noted that it would have been reassuring to know that such positive experiences were possible.

### Current menstruation experiences

Despite often positive initial reactions to menstruation, 87% (104/120) of survey respondents reported ongoing concerns (Figure 1). Universal challenges included heavy periods (48%), pain (45%), practicalities of menstruation (45%), and hormonal impacts on mood and behaviour (40%). Concerns about hormones affecting behaviour, social appropriateness and sleep interruption remained significantly higher among caregivers of adolescents with higher support needs (Table 3).

**Table 3.**
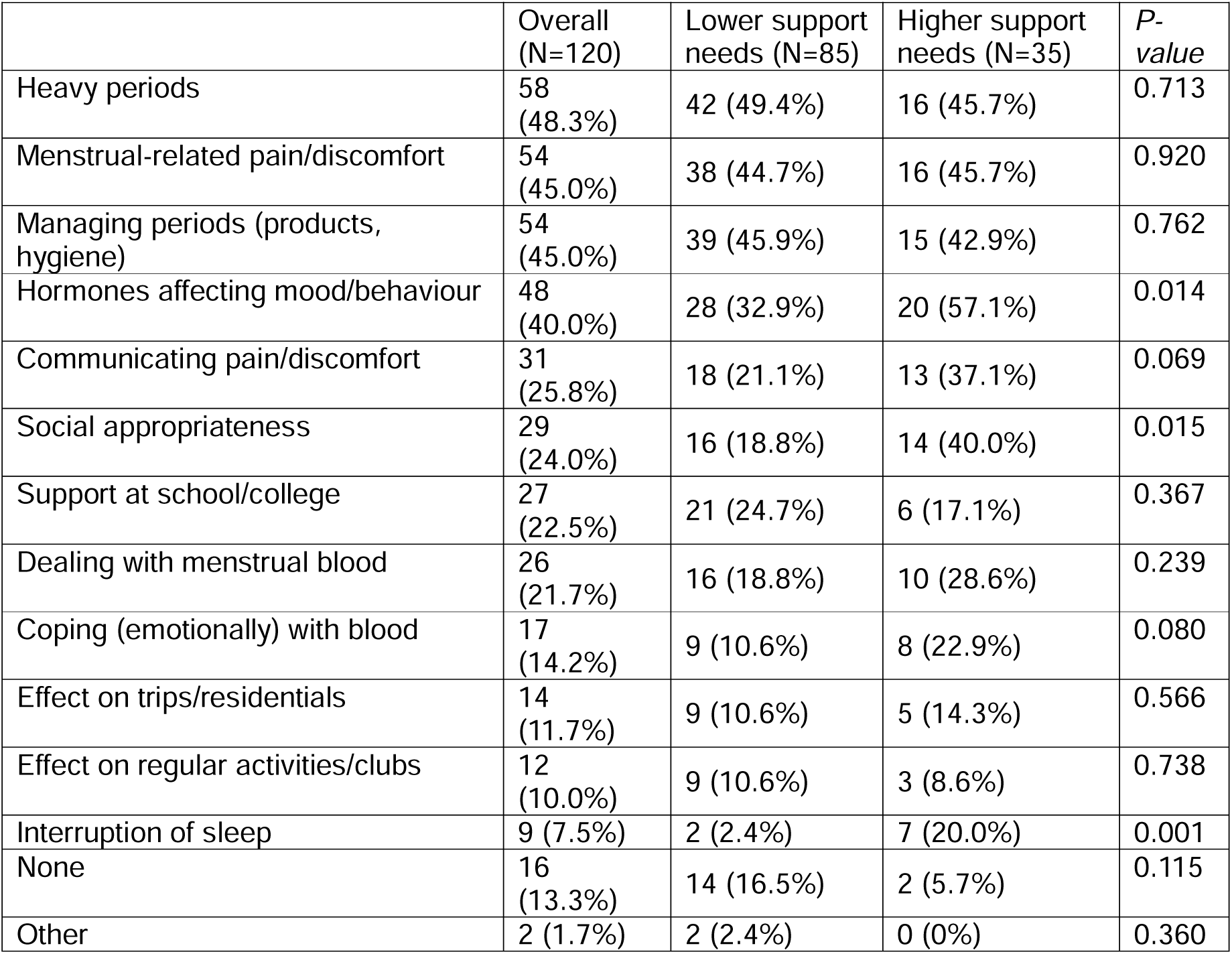
Current caregiver concerns about menstruation by support needs.

#### Understanding of menstruation

Survey data indicated that almost half of adolescents had a good understanding of menstruation (46%, n=66/143) and over a third had some understanding (34%, n=49). Qualitative interviews revealed the variations in understanding. Some adolescents demonstrated a clear grasp of the basic biology, linking the cycle directly to reproduction. For instance, Sofia accurately detailed the physical process and its purpose:

> Researcher: *“And what is a period?”*
>
> Sofia: *“Blood… [from] the vagina.”*
>
> Joanne (Mother): *“And why does it come?”*
>
> Sofia: *“Because your body thinks you’re having a baby… It’s the menstrual cycle.”*

Other caregivers noted that their daughters understood that periods come regularly, but did not appreciate why. Diane shared that her daughter thought she had wet herself every time her period started, while Fiona found that the use of black period underwear made it difficult for her daughter to understand that she was bleeding. Several caregivers expressed concern that their daughter’s physical development was more advanced than their understanding of puberty.

#### Menstrual cycle features and symptoms

Most adolescents (87%, n=104) did not use hormonal medication and complete clinical suppression of the menstrual cycle was rare (3%, n=3). When used, hormonal medication was typically administered by caregivers, who sometimes reported a sense of discomfort if their daughter lacked a clear understanding of medication’s purpose. However, some adolescents had good understanding, as demonstrated by Sienna, who described how her *“special pill”* made her periods less painful and altered her menstrual cycle, stating: *“I don’t have my period every month. My sister has one every month. I have mine every three months.”* Heavy or prolonged bleeding, defined by changing sanitary products every 1–2 hours, periods exceeding seven days, or large blood clots, affected 29% (34/118). Common physical symptoms included menstrual cramps (74%) and fatigue (39%), alongside behavioural symptoms like mood swings (70%), increased irritability (53%), challenging behaviour (30%) and anxiety (24%). One caregiver reported undiagnosed endometriosis (Table S3).

Adolescents generally disliked their periods, frequently describing feelings of anger, distress or sadness, often tied to pain. As Mia explained: *“I don’t like periods that much, because it’s a little bit hard for me, and I don’t like it when I’ve leaked through my bottoms.”* She later clarified that episodes of leakage were rare, but she worried about it. However, Kathryn was adamant that she felt *“happy”* when she had her period. Subsequent discussion with her mother, Julie, suggested that Kathryn may have responded in this way because having her period meant that Kathryn got to spend more dedicated time with her key person at school.

#### Pain and communication

Communicating and managing pain were ongoing concerns for 71% (n=85) of survey respondents (Table 4), with period pain reported by 89% (105/118) of caregivers (Table S3). Adolescents frequently equated pain with periods, as shared by Sofia: *“What I mean by yucky is pain… It’s a horrible thing”*(. Sofia also internalised and normalised this discomfort: *“I try and be brave and just do my work.”*

**Table 4.**
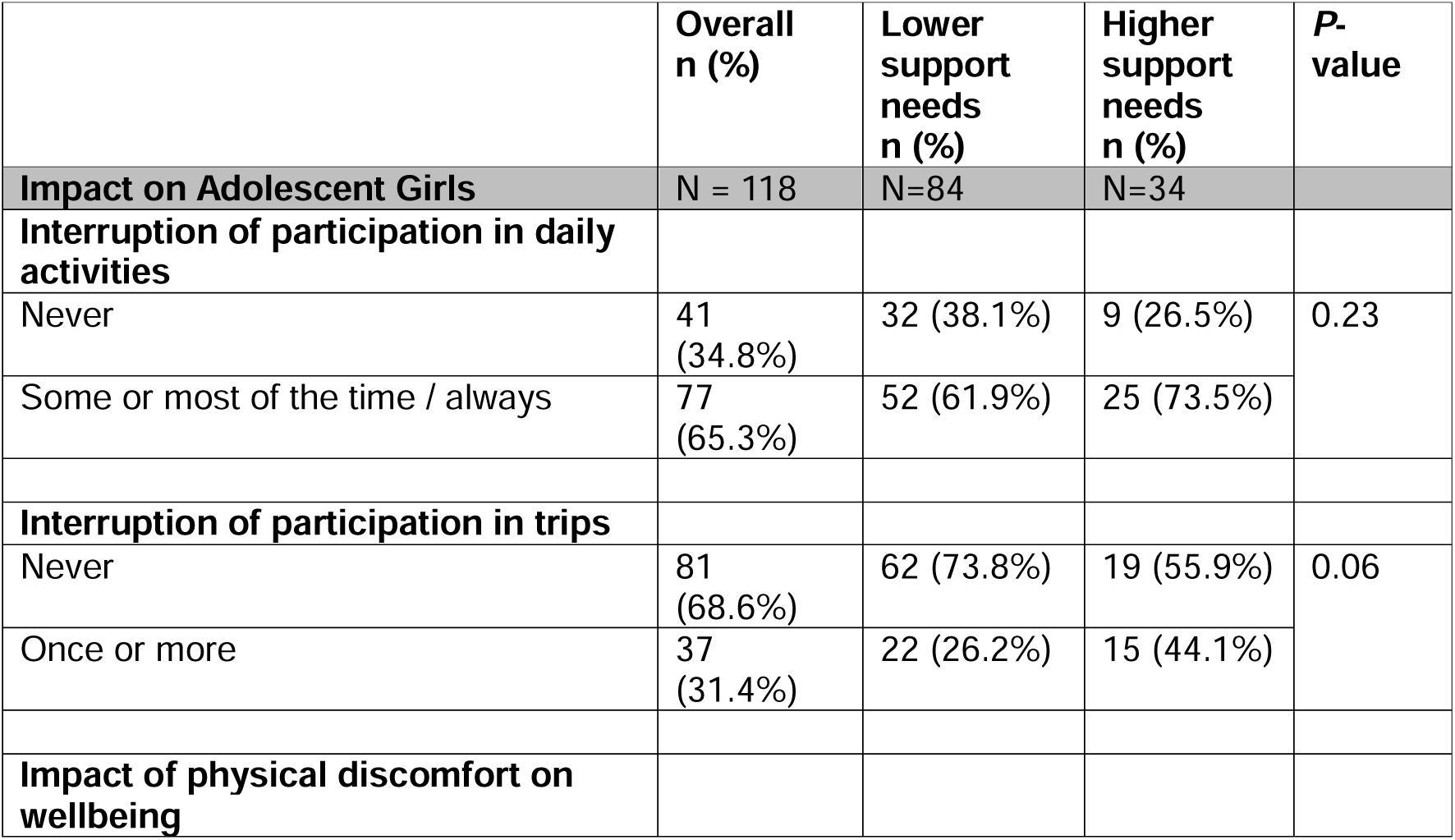
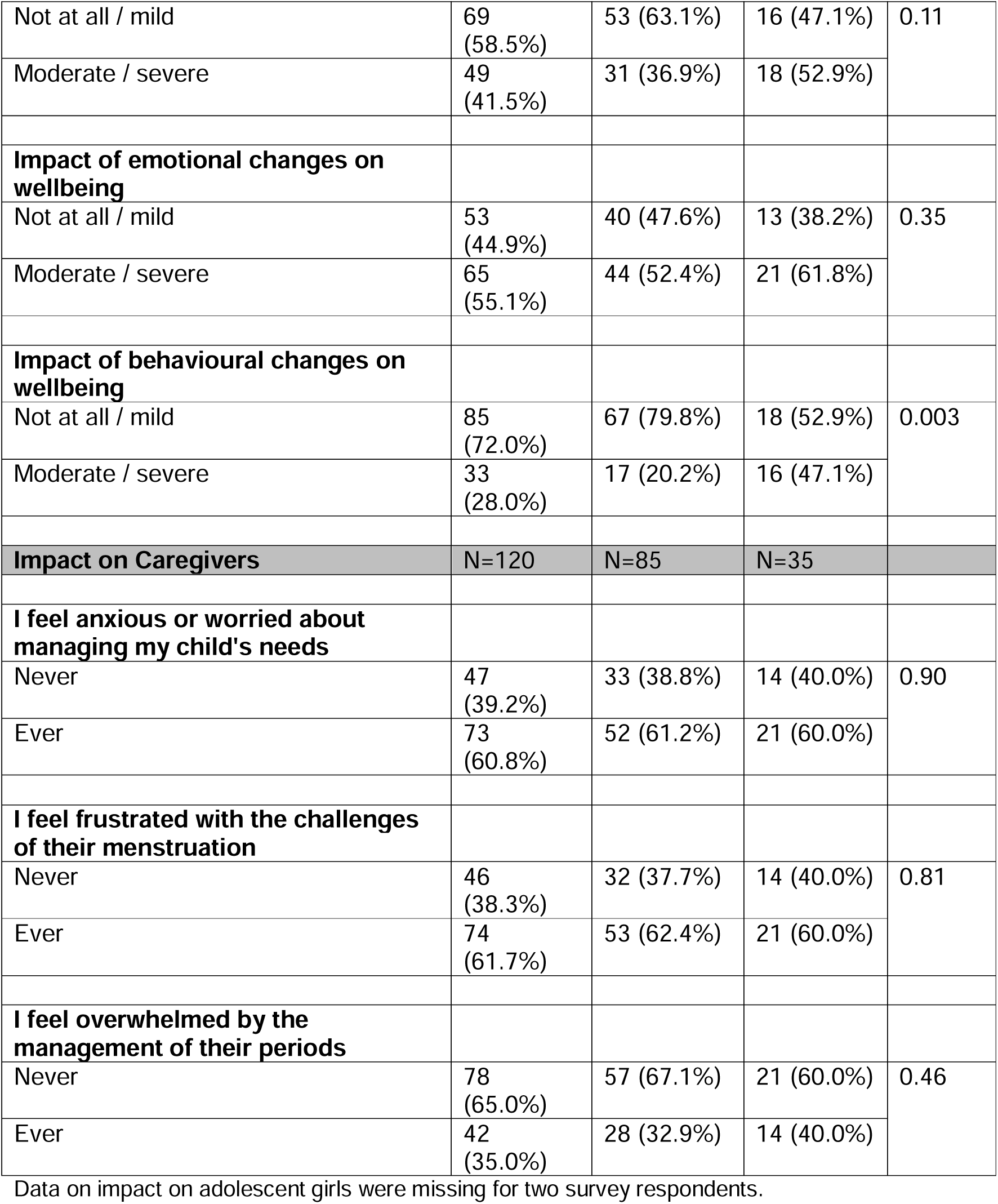
Impact of periods on participation and wellbeing of adolescent girls and their caregivers.

|  | Overall<br>n (%) | Lower<br>support<br>needs<br>n (%) | Higher<br>support<br>needs<br>n (%) | P-value |
| --- | --- | --- | --- | --- |
| **Impact on Adolescent Girls** | N = 118 | N=84 | N=34 |  |
| **Interruption of participation in daily activities** |  |  |  |  |
| Never | 41<br>(34.8%) | 32 (38.1%) | 9 (26.5%) | 0.23 |
| Some or most of the time / always | 77<br>(65.3%) | 52 (61.9%) | 25 (73.5%) |  |
| **Interruption of participation in trips** |  |  |  |  |
| Never | 81<br>(68.6%) | 62 (73.8%) | 19 (55.9%) | 0.06 |
| Once or more | 37<br>(31.4%) | 22 (26.2%) | 15 (44.1%) |  |
| **Impact of physical discomfort on wellbeing** |  |  |  |  |
| Not at all / mild | 69<br>(58.5%) | 53 (63.1%) | 16 (47.1%) | 0.11 |
| Moderate / severe | 49<br>(41.5%) | 31 (36.9%) | 18 (52.9%) |  |
| **Impact of emotional changes on wellbeing** |  |  |  |  |
| Not at all / mild | 53<br>(44.9%) | 40 (47.6%) | 13 (38.2%) | 0.35 |
| Moderate / severe | 65<br>(55.1%) | 44 (52.4%) | 21 (61.8%) |  |
| **Impact of behavioural changes on wellbeing** |  |  |  |  |
| Not at all / mild | 85<br>(72.0%) | 67 (79.8%) | 18 (52.9%) | 0.003 |
| Moderate / severe | 33<br>(28.0%) | 17 (20.2%) | 16 (47.1%) |  |
| **Impact on Caregivers** | **N=120** | **N=85** | **N=35** |  |
| **I feel anxious or worried about managing my child's needs** |  |  |  |  |
| Never | 47<br>(39.2%) | 33 (38.8%) | 14 (40.0%) | 0.90 |
| Ever | 73<br>(60.8%) | 52 (61.2%) | 21 (60.0%) |  |
| **I feel frustrated with the challenges of their menstruation** |  |  |  |  |
| Never | 46<br>(38.3%) | 32 (37.7%) | 14 (40.0%) | 0.81 |
| Ever | 74<br>(61.7%) | 53 (62.4%) | 21 (60.0%) |  |
| **I feel overwhelmed by the management of their periods** |  |  |  |  |
| Never | 78<br>(65.0%) | 57 (67.1%) | 21 (60.0%) | 0.46 |
| Ever | 42<br>(35.0%) | 28 (32.9%) | 14 (40.0%) |  |

Data on impact on adolescent girls were missing for two survey respondents.

Communication barriers were substantial; 31% (37/120) could not verbalise pain, and 12% (14/120) could not communicate pain using words or gestures (Table S3). During interview, caregivers described how they relied heavily on physical signs of distress (e.g., interrupted sleep), intuition and behavioural cues. Emma said: “*She doesn’t want to dance and that’s when we know that there is pain there”*. Pain may also present atypically, for example as *“peals of laughter”,* or be misattributed to the adolescent’s disability: “*What it [period pain] looks like in our people with Down syndrome can look like behaviour”* (Anna, Educator). Several mothers became upset when talking about the challenge of accurately identifying pain: *“She will come looking for cuddles… she’s distressed. She’s got tummy ache. She doesn’t know why, she doesn’t understand.”* (Nicole). Julie also noted: *“The managing the period itself is… not onerous… But it can have an impact, that pain. It’s hard to watch”*.

Pain management primarily involved over-the-counter medication (92/105; 88%) and heat pads (56/105; 53%). Hormonal interventions (10%) and prescription-only medications like naproxen and mefenamic acid (5%) were less common. Twenty-four percent of 105 survey respondents felt pain was not effectively managed.

#### Preparing for and tracking periods

There was a mix of approaches when it came to preparing for upcoming cycles. Some caregivers actively told their daughters that their periods would be coming “soon” or in a specified number of days to help them prepare. However, other caregivers chose to manage the materials and preparations entirely themselves because they felt their daughters did not understand the concept of time.

Many respondents reported that they tracked menstrual cycles using paper calendars or notes, while 29% (35/120) reported using a tracking app: seven adolescents used the app independently or with indirect support, seven used it together with their caregiver, and 22 caregivers used the app themselves. While caregivers often held the responsibility for tracking, adolescents sometimes displayed more independence than realised. During her interview, Sofia surprised her mother by sharing and demonstrating how she managed this task herself: Joanne (Mother): “*We don’t track.”* Sofia: “*I got that on my phone. I track with my phone.”*

#### Social appropriateness

Caregivers and professionals frequently noted that some adolescents lacked a social filter, leading to public disclosures of their menstrual status that they feared could cause discomfort:

> *“She’s quite happy to announce to the whole world she’s got her period. She doesn’t really have a filter on stuff like that.”* (Teacher)

Julie recalled her daughter announcing, *“I have to go and change my pants,”* in front of visitors, commenting that *“It’s understanding what you should share and shouldn’t share.”*

Other adolescents appeared to have internalised messaging about expected norms placed on women and girls to mask menstruation and symptoms. For example, Sofia explained that it is more “polite” to say your tummy hurts than to say that you have your period.

#### Practicalities of menstruation and product choice

The practicalities of menstruation—encompassing product use and personal care—were a concern for 45% (n=54) of caregivers (Table 3). Sixteen percent (19/118) of young people were unaware of the need to manage their period, requiring comprehensive menstruation support (Table S4). Among those with awareness (n=99), 74% always informed their caregiver when their period started. Given that a high proportion of adolescents required physical assistance with products and personal care at home and at school (Table S4), not sharing this information complicated management for caregivers.

The physical impact of providing support was often greater for families of adolescents with more complex needs. Julie articulated the challenges she faces supporting her daughter’s menstruation when they are away from home:

> *“It’s probably the hard part of caring… the physical, personal care side of it. At home - it’s fine. When we’re out and about that’s another thing altogether… standing her up in a bathroom when she’s got her period… it’s just not ideal. You want to be able to lie her down and clean her properly and change her comfortably.”*

The most used menstrual absorbents were period underwear (77%, 89/115) and disposable pads (68%, 78/115), often in combination on heavy flow days to prevent leakage and to simplify management (as a pad could be removed from period underwear partway through the school day to leave a clean layer beneath). Interviewees explained that some adolescents liked wearing pantyliners every day, a practice caregivers supported both to manage vaginal discharge and to provide a safety net for the unpredictable onset of a cycle. Period swimwear was used by 34% (n=39) of adolescents when the flow was light. Only one survey respondent reported tampon use. Menstrual care could be complicated by co-occurring incontinence (19%, 23/119), which necessitated the layering of menstrual products with continence aids and night-time nappies.

Overall, 67% of 70 adolescents were reportedly comfortable using period products, with no significant differences by support needs (*P*=0.541). While pads with adhesive ‘wings’ were described during interviews as *“fiddly*” and unpleasant because they could stick to the skin, 67% of 99 adolescents with menstrual awareness were reported to be moderately or very confident in managing their period in different settings (Table S4). Diane stressed that independence was increasing slowly with time and repetition: *“[She] is a visual learner… I think it’s just keeping the same consistency. So it’s doing the same thing… talking her through the process whilst I’m doing it. It is this repetition [and] you have to be very patient as well.”*

Period underwear was often described by caregivers interviewed as *“magic pants”* or a *“game changer”*, valued for longevity (they often required no change during the school day) and privacy. One mother sought specific brands with logos on the outside to help her daughter identify the inside, enabling her to ensure absorbency and put them on independently. However, their absorbent nature made monitoring flow difficult for caregivers and was occasionally thought to hinder progress towards independence, as the lack of visual blood meant the adolescent was less aware of when a change was required. Rinsing the garments generally fell to the primary caregiver.

Product affordability was a barrier for 11% (13/115) of families, although many considered the cost of reusable period underwear worth it, as Georgina noted, you *“get what you pay for”*, as cheaper versions they had trialled had leaked.

#### Impact on adolescents’ participation and wellbeing

Menstruation affected daily life and overall wellbeing, particularly for those with higher support needs (Table 4). While caregivers often focused on the logistical “worry” of leaking, adolescents themselves identified physical pain as the reason their daily activities were interrupted by their period. Sienna shared: “*When I get a lot of pain I have to stop helping mum in the kitchen. I relax and watch TV.”*

Overall, 65% (77/118) of adolescents experienced interruption to daily activities during menstruation. While most interruptions were occasional (mainly affecting swimming), 21% (7/34) of those with higher support needs were unable to participate in daily activities most or all of the time. Nearly a third (31%, 37/118) had missed trips (Table 4). Caregivers noted during interview that the disruption would have been greater in the absence of period underwear. Caregivers reported moderate-to-severe impacts on adolescent wellbeing due to emotional changes (55%, 65/118) and physical discomfort (42%, 49/118), with no significant differences according to level of support needs. Forty-seven percent of adolescents with higher support needs experienced behavioural changes that had a moderate or severe impact on wellbeing (20% lower support needs, *P*=0.003) (Table 4).

#### Impact on caregiver wellbeing

Although 90% of 120 caregivers felt moderately-very confident providing practical support with menstrual care, anxiety (61%, 73/120) and frustration (62%, 74/120) at the challenges they or their child faced were common, regardless of the adolescent’s support needs (Table 4). Thirty-five percent (42/120) of caregivers felt “overwhelmed” at least some of the time.

Interviewees described a heavy mental load involving cycle prediction and anticipating and managing emotions and household tensions. This mental load could be compounded by a sense of isolation and the “*unavoidable”* role the caregiver needed to play in supporting with menstrual care. As Emma explained: *“There isn’t anyone else I can pass it to… so it’s me or nobody… It has impacted my work; it’s impacted what I’m able to do at weekends”*.

Caregiver distress was also caused by perceived powerlessness to identify or effectively manage pain. Julie noted: “*It’s all a matter of interpreting, so there may be elements of projection, and that’s what I sometimes worry about. I’m not always sure that I’m getting it right”*. While hormonal medication sometimes provided *“peace”* by controlling symptoms and regulating cycles (confirming the absence of pregnancy), it introduced new worries and inner conflict as caregivers weighed benefits against potential long-term health impacts. Future concerns included the adolescent’s eventual level of independence and sources of support in settings like college, as well as sexual activity, pregnancy and the balance of autonomy:

> *“In so many ways she’s growing up. You know, she’s got a boyfriend. She’s desperate to get married, all this sort of thing, yet she’s so immature in other ways. So it’s that whole sort of two sides to it. And you know, that’s…. she’s got a period. She is growing up, but yet she’s, you know, not able to do it by herself…. Not able to look after herself in that respect”* (Fiona).
>
> *“[She] talked at times about ‘I really want to have a baby’, but we just said you can’t… we’ve talked about that’s a very big responsibility… and also the importance of [adolescent] having a choice and how we guide that. Because how much are we making that decision for her if we say you’re not having a baby or you’re not going to be having sex”* (Rachel).

### Support systems and information

#### School support

Most caregivers (71%, 85/120) reported that their child received school-based menstruation education, of whom 52% (n=44) found it appropriate. Anna (Educator) noted that mainstream schools do not always provide tailored information: *“I think it varies in our kids experience, because it could be that some schools would just automatically include them in a PSHE [Personal, Social, Health and Economic] lesson, which can be fine and can have value. But they’re not necessarily supplementing it with the stuff that’s accessible and at the right point for that child”*. In contrast, interviewees described special schools as being more versed in the use of accessible formats, including delivery of information on menstruation via videos, social stories and through practical sessions, such as putting sanitary towels in underwear.

Special school settings were also generally considered to be better equipped to integrate menstrual care because they were already providing high levels of support for toileting and personal care for many of their pupils. In contrast, adolescents in mainstream schools had less physical support due to resource availability and safeguarding regulations, with care responsibilities passed back to family during the school day: *“They’re so lovely I know that they would [help], but it’s just that line that you can’t cross, so they have to ring me. And I have to leave work and go and pick her up.”* (Vicky).

#### Medical support and hormonal medication

Concerns about heavy bleeding and pain led 42% (50/120) of caregivers to seek clinical support. Of these, 46% (23/50) were offered hormonal medication and 26% (13/50) were offered non-hormonal options (e.g., tranexamic acid). Over a third (36%) of caregivers were not satisfied with their medical consultations. A common source of this dissatisfaction was perceived reluctance by medical professionals to prescribe hormonal medication due to the young age of the child or the need to observe persistence of symptoms over time. Caregivers understood and appreciated that approach, but felt frustrated about the lack of support for pain relief. Caregivers also described feeling unsupported as they explored options to manage problematic menstruation, feeling that menstrual pain was dismissed as normal, and reporting a lack of evidence-based guidance to help them weigh potential side effects against benefits:

> *“They don’t give you a sense of ‘we would advise you to try this.’ It’s very much ‘oh, you might want to consider this’… so you’re feeling your way all the time”* (Julie).

This lack of perceived direction often left caregivers feeling helpless, particularly when attempting to manage complex co-occurring conditions like Hidradenitis suppurativa, a chronic inflammatory skin condition that is significantly more prevalent and presents earlier in people with Down syndrome (30).

Conversely, Tracey (GP) explained that there is no age restriction on hormonal treatment and emphasised her commitment to *“individually tailored conversations*” that consider the adolescent’s broader health profile and the family’s preferences.

Reasons for using hormonal medication described in interviews varied. Beyond pain relief, the primary drivers were the desire to regulate or “skip” periods for practical reasons (e.g., holidays). Hormonal suppression also served as a protective measure, with the decision to go on the pill described by one caregiver as a response to fears over a late period:

> *“I just had this freak out one time when her period was late. And I just thought, what if someone’s abused [her]… my head was spinning. So I did think, you know what, with all the other reasons to go on the pill, then that was the decision I made. And the GP was very supportive of it. And the other side of going on the pill is that since, when she does have her period, it’s more manageable – much lighter and less pain”* (Georgina).

#### Information demand

Overall, 26% (6/23) of caregivers of adolescents who had not yet started menstruating and 53% (63/120) of those whose children had started menstruating sought support to prepare for the onset of periods. Information was primarily obtained from online resources (55%), books (52%), and other parents of children with Down syndrome (35%); medical professionals, charities, teachers and other sources were drawn on to a lesser extent. Although the majority (83%, 57/143) were able to locate the information they sought, resources may not be widely disseminated in clinical communities.

Almost all survey respondents expressed a demand for further support (Table S5). Priorities included: 1) intellectual disability-specific resources for young people, covering the biology of periods and the menstrual cycle in an accessible format; 2) practical resources to support independent management of periods; and 3) pain management strategies, including guidance on identifying and alleviating distress in those with communication barriers. Caregivers requested that resources be easy to locate and simple to digest. They also asked for information on how periods impact people with Down syndrome and guidance on managing complex needs.

While medical professionals viewed the management of comorbidities like cardiac or renal conditions as a standard part of individualised clinical assessment, they identified a significant gap in tailored resources. Tracey (GP) noted that while general ‘Easy Read’ guidance exists, there is a lack of accessible information for families regarding the safety and suitability of medical options in the context of the complex health profiles that can be associated with Down syndrome. Shortage of staff trained to provide menstrual support for people with intellectual disabilities was also noted by several professionals.

## Discussion

To our knowledge, this study represents the first comprehensive mixed-methods investigation into the menstrual health of adolescents with Down syndrome in the UK to integrate quantitative survey data alongside the direct voice of young people and the perspectives of family and professional caregivers. We have documented considerable variation in the awareness and understanding of menstruation, and the degree of practical support required for safe and dignified menstruation. Our findings suggest that while adolescents displayed a range of reactions to menarche, their overall response was often much better than their caregivers had anticipated; crucially, our data highlight that the widespread adoption of period underwear significantly simplified menstrual care routines, aiding adolescent dignity and enabling more independent self-care. Despite this, menstrual cycles impact both adolescent and caregiver wellbeing. Key challenges pertained to the communication and management of pain, the impact of hormonal changes on mood and behaviour, and the complexities of menstrual care: challenges which “overwhelmed” one in three primary caregivers and can greatly impact family wellbeing (10, 26).

The median age of menarche in our sample was 12, aligning with earlier UK (21) and international data for people with Down syndrome (31, 32) and the general UK population (33). A critical takeaway is that menstruation education must be appropriately tailored to an individual’s level of understanding and emotional maturity, which often lags behind biological development. As reported by a previous study (21), many caregivers, and even some healthcare professionals, frequently assume that menarche will be delayed in line with other developmental milestones. Although we also observed this assumption, fear of early menarche and its consequences was also reported in our study. Caregivers should be advised that menarche typically occurs within the standard population range to correct misinformation and alleviate fears of precocious puberty. Although evidence remains limited and inconsistent, some studies suggest that early menarche may limit final adult height (34). Providing families with accurate information is essential to mitigate unnecessary distress.

Research has shown that when young women with intellectual disabilities are unprepared for menstruation, menarche is often experienced as a frightening or distressing “shock” (21, 35). In our study, reactions to menarche were varied, but advance preparation was not associated with more positive responses. The discrepancy between a caregiver’s anxiety before menarche and their child’s actual response suggests that caregivers often anticipate a worst-case scenario, potentially because society frequently frames menstruation as an unmanageable hygiene crisis or a major burden (8, 36). The reality of menstruation was not easy—the adolescents in our study still experienced fear and worry during menarche, and ongoing distress, as evidenced by the 91% of caregivers reporting persistent concerns about menstruation. However, the onset of periods was ultimately a manageable milestone, and not the crisis that caregivers had feared.

There is a lack of quantitative survey data with which to compare our findings. The qualitative study conducted by Mason and Cunningham (2008) reported that around one-third of women with Down syndrome could achieve independent menstrual self-care, with an additional third capable of self-care with the support of reminders (21). Although not directly comparable due to differences in questions asked, in our study two-thirds of adolescents with menstrual awareness were reported to feel confident managing their periods across different settings, and demonstrated high levels of autonomy (particularly at school). In contrast with Mason and Cunningham, who noted low parental expectations regarding autonomy, we found caregivers and adolescents mutually desired and pursued independence. This is exemplified by Sofia independently tracking her cycles on her phone without her mother’s knowledge. This shift in expectations and behaviours may reflect both the long-term impact of early intervention frameworks, and movement away from deficit-focussed narratives. This is reflected in recent calls for healthcare and educational models that actively prioritise autonomy, identity, and active participation for individuals with Down syndrome (37, 38). In addition, the introduction of period underwear has vastly simplified practical menstrual care, reducing both the physical and sensory barriers associated with some other products. Our data strongly reinforces the notion that with appropriate targeted training and visual aids, many young people can achieve substantial, dignified autonomy in their menstrual care (15, 21, 39).

A previous clinical review article reported that cyclical hormonal changes are associated with a higher prevalence of behavioural change in people with intellectual disabilities than in the general population (16). Our survey findings align with this, showing high overall caregiver concern regarding changes in behaviour, particularly for adolescents with higher support needs. We did not fully explore the specific nature of behavioural symptoms and their consequences in this study. However, others have reported presentations including self-injury and aggression (16, 25). This may be driven in part by an inability to articulate menstrual pain.

We also found indications of the gendered construction of pain, which refers to the way pain is socially produced, interpreted, and legitimised through gendered norms (40), as evidenced by Sofia’s report: *“I try and be brave and just do my work.”* Menstrual pain (dysmenorrhea) is commonly trivialised, normalised and framed as a routine aspect of womanhood, something that should be endured and a private issue, rather than a public health issue (41). This cultural expectation of “getting on with it” is linked to stigma-based societal norms of “menstrual etiquette”, which mandate total containment and concealment of periods. As Wiggleton-Little states, simply sharing experiences of menstrual pain can violate this etiquette and result in social sanctions (e.g., criticism and exclusion) (42). Consequently, silence and the culture of secrecy, driven by menstrual stigma, make experiences of painful periods less visible and more likely to be overlooked or dismissed. For individuals with communication or cognitive difficulties, navigating these social expectations may be particularly confusing and overwhelming, or they may simply not be aware of them. Further, the systemic normalisation of pain is particularly concerning given that heavy (menorrhagia) and painful periods are prevalent in our study, as in earlier work (20, 21). This burden is compounded by common co-occurring physical conditions; for example, the cyclical hormonal “flare-ups” associated with Hidradenitis Suppurativa can complicate menstrual care and substantially impact quality of life (43). Overall, there appears to be an evidence gap concerning communication abilities and presentation of premenstrual and menstrual symptoms, including dysmenorrhea, in people with intellectual disabilities, which could be relevant for intervention. For caregivers, menstruation can raise complex tensions between supporting the adolescent’s developing autonomy and agency, and managing concerns about reproductive vulnerability, potential risks, and the practical demands of care. This emotional conflict between honouring a young person’s journey to adulthood and the need to protect them aligns with the wider literature on intellectual disability and sexuality (44). Our findings emphasise that this remains a relatively neglected area for families of adolescents with Down syndrome. In our study, caregivers felt they lacked guidance on how to navigate conversations and decisions around physical maturity, variable communication skills, and social openness.

This lack of guidance is particularly evident when navigating the clinical management of menstruation. In our sample, one in five adolescents had been offered hormonal interventions, and 13% were currently utilising them for symptom management or cycle suppression. This rate of use mirrors the wider UK general population (45), but is lower than reports concerning the broader intellectual disability population, where hormonal suppression has traditionally been more prevalent (11, 46). While hormonal options can effectively alleviate pain and heavy bleeding (47), their use raises ethical considerations regarding autonomy, and requires careful balancing of potential side effects and long-term impacts (23, 26, 31, 48). It is therefore unsurprising that caregivers in our study felt a heavy sense of responsibility regarding this choice. To mitigate this burden, there is a clear need for developmentally appropriate, Easy Read decision aids that facilitate adolescents to meaningfully participate in these conversations alongside their families when possible. There is also a corresponding need for clear, evidence-based guidance for clinicians to facilitate clinical consultations (48).

Our findings argue for a shift from a deficit-based management model toward a holistic framework of “menstrual wellbeing” for adolescents with Down syndrome. True wellbeing in this population involves physical comfort, peace of mind, communication agency, and bodily dignity. Currently, the wellbeing of many young people is heavily compromised by communication barriers. Integrating menstrual and somatic symbols into Augmentative and Alternative Communication (AAC) systems will give some young people the vocabulary to discuss menstruation. There is also a need to co-develop accessible visual tracking tools, paired with further research into pain identification and management. Furthermore, menstrual wellbeing requires communication between adolescents, caregivers, and schools. Reusable period underwear represent a vital tool for preserving privacy and dignity within this wider support structure. Ultimately, achieving true menstrual wellbeing demands co-developed support systems that span educational, social and clinical spaces, including frameworks to track menstrual cycles and symptoms, tools to aid communication of menstrual pain and needs, caregiver and adolescent peer networks, and accessible clinical decision aids.

### Implications for Policy and Practice

Our findings identify a high demand for resources and indicate unmet needs that should be addressed. While some needs can be addressed through better signposting to existing materials, tailored interventions are also required. We propose four key pillars for future support: 1) Tailored educational resources, 2) Guidance and resources on practical menstrual care 3) Standardised clinical training and tools and 4) Creation of supportive community environments. Figure 2 presents these pillars in more detail, showing how each was informed by findings from our study.

**Figure 2.**
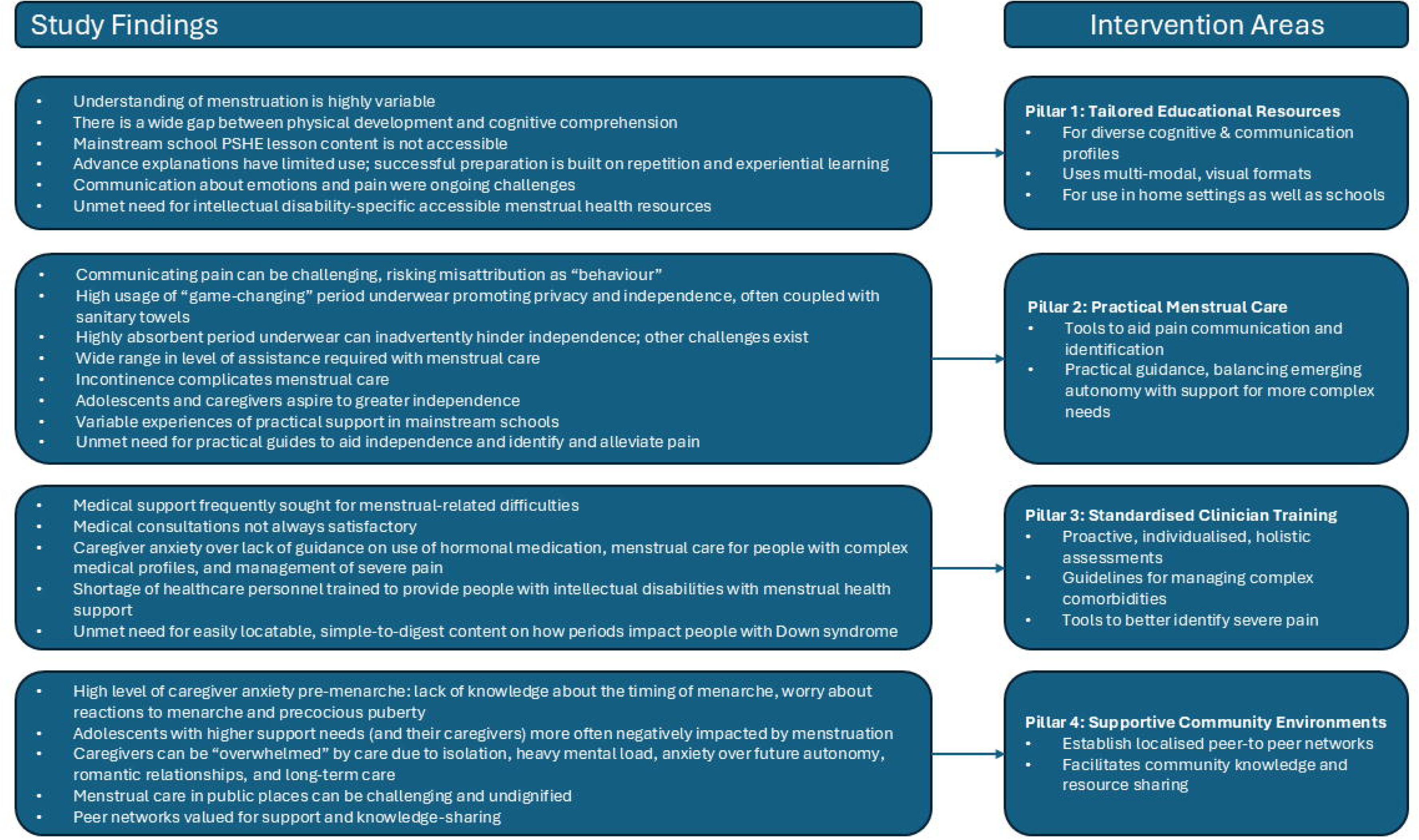
Inductive framework showing how study findings (left) map onto the proposed intervention pillars (right)

The findings of this study are broadly consistent with the recommendations of previous research (10, 11). While these suggestions are based on the lived experiences of our participants, any resulting recommendations or interventions must be co-developed with young people with Down syndrome and their families and professional supporters. The feasibility and acceptability of any resulting interventions should also be rigorously assessed across diverse settings before widespread implementation.

### Strengths and Limitations

A major strength of this study is the mixed-methods approach, which successfully integrated caregiver perspectives and the direct voices of adolescents, a population that is traditionally excluded from research, alongside a national survey of caregivers. This allowed for both statistical breadth and qualitative nuance across a range of cognitive abilities and educational settings.

We must not diminish the value of the direct adolescent contributions. Our final sample size comprised six adolescents with Down syndrome. Broader recruitment was constrained by logistical challenges of scheduling face-to-face interviews around full-time education, decisions of some adolescents not to participate, and cases where individuals lacked the capacity to consent. Nevertheless, our qualitative adolescent sample size was comparable to those reported in other studies exploring menstrual health among women and girls with learning or communication impairments, including those with Down syndrome, where sample sizes ranged from 2 to 11 (21, 49, 50). Crucially, these face-to-face adolescent interviews provided rich contextual data and generated insights that would not have been gained from proxy responses alone are therefore a strength of this study. For participants with limited verbal communication skills, their physical conduct, non-verbal expressions of comfort or discomfort when interacting with menstrual products, and their engagement with the visual aids formed important primary data. As an individual’s menstrual experiences are so closely supported by their family, the predominance of caregiver data in this study simply reflects how young people in this study are supported in their everyday lives.

As the Down syndrome community is very diverse in cognitive and physical ability, many of the findings and potential strategies for intervention are relevant to the wider intellectual disability community.

However, several limitations must be noted. First, our small qualitative sample of six adolescents living in London and neighbouring counties limits the range and diversity of experiences that were captured, and thus the transferability of the findings to the wider population of adolescents with Down syndrome, particularly those with more complex cognitive and communication needs, or those from different ethnic and geographical backgrounds. Second, while the survey was advertised extensively, recruitment via social media and Down syndrome groups means the experiences of families who do not use social media and do not engage with support services are not captured. This demographic may face additional menstruation barriers if they represent a more isolated population. Third, while the survey achieved broad geographical representation across the UK, the sample lacked ethnic diversity, with 86% of survey respondents identifying their daughter as White. Our findings therefore do not fully capture the diverse cultural and religious factors that likely shape how menstruation is perceived, experienced and discussed within minority ethnic communities, or how these factors uniquely intersect with intellectual disability. Future research must target underrepresented and underserved groups and intentionally employ an intersectional lens to explore how the overlaps of race, gender and disability shape menstrual health experiences and compound health inequities in the UK.

Additionally, we cannot exclude the possibility that self-selection into the study may have produced a sample skewed toward either highly engaged caregivers with strong support systems, or conversely, caregivers motivated to respond due to the severity of their child’s negative menstruation experiences. While this potential bias could reduce generalisability of the quantitative data to the broader UK Down syndrome population, as a wide spectrum of lived experience was reported in both the quantitative and qualitative components of the study, we do not believe that self-selection compromised the overall interpretation of our findings. As caregiver assessment of the level of assistance required for personal care was subjective, it is possible that the support needs of some adolescents were mis-categorised. Subgroup analyses should be interpreted with caution due to the small sample size.

Furthermore, the qualitative interviews were restricted to London and the surrounding counties, which may not fully reflect the economic diversity or the variations in healthcare access and support services available across the rest of the UK. Finally, while we identified caregiver concerns regarding behavioural changes, our study did not fully explore the specific nature of these symptoms, nor were we able to quantify menstrual pain; these remain areas for future inquiry.

## Conclusions

Menstrual health for adolescents with Down syndrome is a highly individualised experience, shaped by specific cognitive and physical support needs. Our study highlights that achieving positive menstrual health outcomes requires equipping caregivers with the tools to support their young people effectively, alongside accessible and appropriate guidance to foster young people’s independence, choice and autonomy. Addressing unmet needs through accessible information, practical menstrual care resources, and professional guidance will facilitate safe and dignified menstruation.

## Data Availability

The data presented in this paper are hosted in the London School of Hygiene & Tropical Medicine data repository at https://doi.org/10.17037/DATA.00005282. Anonymised data will be made available for use in ethically approved research by completing the application form.
Due to the sensitive nature of the interviews and the potential for participant identification within a small, specific community, qualitative transcripts are not publicly available.

## Acknowledgements

We are indebted to all of our Advisory Group, especially the mothers and teenagers with Down syndrome who shaped the research approach and study tools through their expert lived experience. We are also immensely grateful to all of our study participants for taking the time to share their personal experiences. Special thanks also to Kat Booker for translating the survey into Welsh, and to Vishna Shah at the London School of Hygiene & Tropical Medicine for her help coding transcripts from the interviews with professionals. We would also like to thank Down Syndrome UK, Down Syndrome Scotland and the Down’s Syndrome Association, along with all the local Down syndrome organisations across the country who disseminated the survey to their members, and in some cases also identified eligible interview participants. Finally, a huge thank you to the Down’s Syndrome Research Foundation (DSRF) UK, without whose financial support this research would not have been possible.

## Supporting information captions

**Supplementary Table 1. Conceptual Framework and Associated Research Questions**

**Supplementary Table 2. Qualitative Codebook for Caregiver, Adolescent and Professional Interviews**

**Supplementary Table 3. Features of the menstrual cycle and premenstrual symptoms**

**Supplementary Table 4. Level of support required to manage menstruation**

**Supplemental Table 5. Areas of demand for menstruation support**

