## Supplemental Tables for "Navigating Menstruation in Adolescents with Down Syndrome: A Mixed-Methods Study of Caregiver Reflections and Adolescent Voices in the UK"

**Supplementary Table 1. Conceptual Framework and Associated Research Questions**

| Socio-ecological framework for MH, adapted for disability (39) | | Integrated model of menstrual experience (9) | | Research Areas |
| --- | --- | --- | --- | --- |
| Factors that support MH | **Sub-factors** | **Antecedents** |  |  |
| Societal and government policy factors | Policies, strategies and curriculum; training standards and practices; traditional norms, practices and cultural beliefs | Socio-cultural context; menstrual stigma, gender norms | Behavioural expectations: enforced by others and self | Openness of comms around menstruation. Is it secret? Is infomation provided? What information is provided? In what format? Is it understandable? (relevant for adolescents and caregivers) |
| Environmental and resource availability factors | Water and sanitation facilities including for solid waste management; availability of affordable, usable and culturally appropriate sanitary protection materials | Resource limitations | Physical environment: water, sanitation facilities; economic environment: product affordability and availability | What materials are used? Satisfaction levels? Comfort, affordability, effectiveness? Who makes decisions about what materials to use? Factors that affect the choice of materials used? E.g. educational setting, incontinence, preference of adolescent/caregiver/school, care needs, time since menarche |
| Interpersonal factors – *person with disabilities* | Relationship with family, caregiver (family and / or professional); relationships with healthcare workers, teachers and other people in authority; relationships with peers; perceptions of changes in gender roles post-menarche | Socio-cultural context: menstrual stigma, gender norms | Social support: family, friends, teachers, healthcare workers; behavioural expectations: enforced by others and self | Communication (what? to whom?) during and about menstruation. Institutional setting (school type), Speech and Language Therapists, other input, menstruating female siblings, caregivers |
| Interpersonal factors – *Caregiver* | Relationship with family, the person with disabilities; relationships with healthcare workers and other people in authority; relationships with the wider community; perceptions of changes in gender roles post-menarche |  |  | Prior experiences/ behaviours as a caregiver (overly protective/risk adversity). Relationship with person with Down syndrome before and after menarche. Networks (connectedness/support) and their focus / age ranges of children in groups. |
| Personal factors *– person with disabilities* | Knowledge about the biology of menstruation and menstrual health, information on menstruation and menstrual health; skills in coping and behavioural adaptions (including pain relief); attitudes, beliefs and feelings about menstruation (including sterilisation / long-term contraception); ability to manage menstruation independently, and support required |  |  | Person with Down syndrome: understanding of menarche /menstruation and how to manage? Attitudes, beliefs, feelings about menarche /menstruation. Ability to express pain, feelings. Confidence, capability, comfort, preparedness. Dexterity. |
| Personal factors – *Caregiver* | Knowledge about the biology of menstruation and menstrual health, information on menstruation and menstrual health; skills in coping and behavioural adaptions (including pain relief); attitudes, beliefs and feelings about menstruation (including sterilisation / long-term contraception); ability to manage another person's menstruation independently, support required, and caring tasks related to menstrual health |  |  | Attitudes/beliefs/feelings about mensuration for themselves (influencing person with Down syndrome’s attitudes). How they prepared for menarche. How they prepare for menstruation now. Is what they do (or are able to do) in line with their preferences? |
| Biological factors | Menstrual variations due to age and features of menstrual cycle (regular, irregular, heavy, light) and any other biological changes related to menstruation; intensity of menstruation (pain) and influences on behaviour, health and concentration; biological issues that impact on menstrual health, such as incontinence | Socio-cultural context: menstrual stigma, gender norms | Knowledge: menstrual biology, reproduction, accuracy of taboos, practical management | What are the biological factors experienced (fever, vomiting, pain, fainting, endometriosis, hormone changes, constipation – iron medication. Incontinence and menstrual health?  Down syndrome with & without autism and other disabilities |

**Supplementary Table 2. Qualitative Codebook for Caregiver, Adolescent and Professional Interviews**

| Theme | Sub-theme | Description |
| --- | --- | --- |
| **1. MENARCHE** |  | General category capturing early experiences and introduction to menstruation. |
|  | Experience | What happened |
|  | Feelings | How the person felt |
|  | Preparation | Anything done to prepare the person for menstruation |
|  | Quotes | Illustrative participant quotes regarding menarche |
|  | Resources | Materials or guides used during this phase |
|  | Support | Support provided to person or carer to help them prepare and understand what would happen and what was happening. Support provision during pre-pubescence and/or menarche |
| **2. CURRENT SITUATION** |  | Core category capturing menstrual management, challenges, and lived experiences |
|  | **1. Impact & wellbeing** | Overarching sub-theme for emotional and daily life impacts. |
|  | Caregiver Beliefs | Positive, negative, and the source of that, eg accurate or inaccurate information. Taboos and misconceptions. |
|  | Caregiver Feelings | Emotional state and responses of the caregiver. |
|  | Individual Feelings | Emotional state and responses of the individual with Down syndrome. |
|  | Individual Participation | Levels of participation during menstruation |
|  | Quotes | Illustrative quotes regarding impact and wellbeing |
|  | **2. Info & Resources** | Overarching sub-theme for materials and education Core category capturing contemporary menstrual management, challenges, and lived experiences. |
|  | Gaps | Missing information, services, or resource needs |
|  | Useful resources | Materials, tools, or resources found to be helpful |
|  | Quotes | Illustrative quotes regarding information and resources. |
|  | **3. Knowledge & Practices** | Overarching sub-theme for understanding, routines, and behaviours |
|  | 'Inappropriate' behaviours | Anything considered inappropriate by individuals, families or society |
|  | Communication & openness | Openness related to talking about menstruation (including any taboos, secrets, social etiquette) |
|  | Menstrual biology & Understanding of Menstruation | The individual's understanding of menstrual biology and menstruation more broadly. Challenges faced maturing physically but not mentally |
|  | **4. Practical management** | Overarching sub-theme for the day-to-day routine of handling a period |
|  | Hygiene | Regularity of changing the mens product, bathing, showering, handwashing |
|  | Independent tasks | Any tasks person does independently |
|  | Support provided | Provided by carers/families. Wider support provided by schools etc is coded under ‘Social Support’ |
|  | Tracking & Preparing | Any activities undertaken by person and/or carer |
|  | Quotes | Illustrative quotes regarding practical management |
|  | **5. Sex & babies** | Overarching sub-theme regarding reproduction and conversations about babies |
|  | **6. Menstrual Product** | Overarching sub-theme regarding specific management products |
|  | Disposal & Reuse | How products are disposed of or cleaned/reused |
|  | Product Used, Affordability & Availability | Includes levels of (dis)satisfaction and rationale for using the product, if different products are used concurrently or separately and the reason for that. |
|  | Quotes | Illustrative quotes regarding menstrual products |
|  | **7. Menstrual variations** | Overarching sub-theme for physical cycle differences and medical management |
|  | Features of cycle | Cycle length, flow, health, concentration |
|  | Hormonal medication | Any hormonal medication that impacts periods |
|  | Medical support & information | Provided prior to hormonal medication and when taking it |
|  | Rationale | Reason the CG put the person on hormonal medication (eg heavy flow, pain, to regulate etc) |
|  | Incontinence & medical conditions | Any medical conditions that impact the menstrual experience |
|  | **8. PMS (Premenstrual Syndrome)** | Overarching sub-theme for pre-period and baseline symptoms |
|  | Experiences | Physical and emotional symptoms experienced before and during the menstrual period. Symptoms can include pain, mood swings, anger, fatigue, bloating, and breast tenderness. |
|  | Pain relief | Anything taken to relieve pain but also address any other PMS symptoms, including emotional support, cuddles |
|  | Quotes | Illustrative quotes regarding premenstrual syndrome experiences |
|  | **9. Social support** | Overarching sub-theme for support outside the immediate family unit |
|  | Networks | Broader community or organisational support networks |
|  | NHS | Interaction, support, or guidance from National Health Service provisions |
|  | Peers | Peer relationships, interactions, or comparisons regarding periods |
|  | School & College | Specific institutional support or challenges within educational environments, including if they talk about changing their menstrual materials in school/college |
|  | Quotes | Illustrative quotes regarding social and institutional support |
| **3. RECOMMENDATIONS** |  | Recommendations for others and/or how to fill existing resource gaps. |
| **4. WIDER DISABILITY EXPERIENCE** |  | Codes to capture the broader context of living with an intellectual disability, outside of menstruation |
|  | Pregnancy & birth | The impact of knowing or not knowing that their child would be born with Down syndrome |
|  | School experiences | Experiences in mainstream and special schools unrelated to menstruation |

**Supplementary Table 3. Features of the menstrual cycle and premenstrual symptoms**

|  | **n (%)** |
| --- | --- |
| **Period frequency (for those with natural cycles) (N=104)** |  |
| Regular (approximately monthly) | 86 (82.7%) |
| Irregular (varying intervals) | 15 (14.4%) |
| Don’t know | 3 (2.9%) |
| **Bleed frequency (for those on hormonal medication) (N=16)** |  |
| Planned breaks with bleeding | 8 (50.0%) |
| Unpredictable or irregular bleeds | 6 (37.5%) |
| Periods completely suppressed | 2 (12.5%) |
| **Usual period length (N=118)** |  |
| 1 to 2 days | 4 (3.4%) |
| 3 to 5 days | 67 (56.8%) |
| 6 to 7 days | 37 (31.4%) |
| 7 days or more | 5 (4.2%) |
| Varies | 5 (4.2%) |
| **Experiences heavy or prolonged bleeding* (N=118)** |  |
| Yes | 34 (28.8%) |
| No | 84 (71.2%) |
| **Experiences period pain (N=118)** |  |
| Always | 46 (39.0%) |
| Sometimes | 59 (50.0%) |
| Never | 13 (11.0%) |
| **Able to express with words or gestures when in pain (N=120)** |  |
| Yes | 104 (88.1%) |
| No | 14 (11.9%) |
| **Expression of period pain (N=105)** |  |
| Crying or fussiness | 32 (30.5%) |
| Withdrawing from activities | 23 (21.9%) |
| Difficulty sleeping | 13 (12.4%) |
| Changes in appetite | 12 (11.4%) |
| Verbal complaints of pain | 83 (79.0%) |
| Other* | 8 (7.6%) |
| **Physical symptoms associated with periods (N=118)** |  |
| Cramps | 87 (73.7%) |
| Fatigue | 46 (39.0%) |
| Bloating | 35 (29.7%) |
| Bowel symptoms | 30 (25.4%) |
| Headaches | 18 (15.3%) |
| Other | 8 (6.7%) |
| None | 9 (7.6%) |
| **Has endometriosis (N=120)** |  |
| Yes (diagnosed) | 0 |
| Yes (undiagnosed) | 1 (0.8%) |
| No | 73 (60.8%) |
| Don’t know | 46 (38.3%) |
| **Changes in mood or behaviour associated with periods (N=118)** |  |
| Mood swings or emotional outbursts | 82 (69.5%) |
| Increased irritability | 62 (52.5%) |
| Increased challenging behaviour (more than usual ups and downs) | 35 (29.7%) |
| Increased anxiety | 28 (23.7%) |
| Changes in appetite | 20 (16.9%) |
| Difficulties sleeping | 17 (14.4%) |
| Difficulty concentrating | 13 (11.0%) |
| Withdrawal from social activities | 13 (11.0%) |
| Other | 5 (4.2%) |
| None | 14 (11.9%) |

* Possible signs of this might be they have to change their pad or tampon less than every 1 to 2 hours because it is soaked, bleed longer than 7 days, or they pass large clots.

** Other mainly selected because the young person cannot communicate about pain so hard to know/pin point

**Supplementary Table 4. Level of support required to manage menstruation**

|  | **n (%)** |
| --- | --- |
| **Young person informs someone when they start their period** |  |
| Yes | 77 (64.2%) |
| No | 18 (15.0%) |
| Sometimes | 22 (18.3%) |
| Not applicable – periods are completely suppressed with no bleeding | 3 (2.5%) |
| **Level of confidence of young person in managing their period in different settings** |  |
| Not at all confident | 5 (4.2%) |
| Slightly confident | 28 (23.7%) |
| Moderately confident | 42 (35.6%) |
| Very confident | 24 (20.3%) |
| Not applicable – child/young person is unaware of need to manage periods | 19 (16.1%) |
| **Young person can ask for help managing their period if needed** |  |
| Yes | 58 (82.9%) |
| No | 12 (17.1%) |
| **Level of support needed to use menstrual products at home** |  |
| Completely independent | 7 (10.0%) |
| Indirect support | 15 (21.4%) |
| Partially assisted | 32 (45.7%) |
| Fully assisted | 16 (22.9%) |
| **Level of support needed to use menstrual products at school/college** |  |
| Completely independent | 19 (27.1%) |
| Indirect support | 19 (27.1%) |
| Partially assisted | 22 (31.4%) |
| Fully assisted | 10 (14.3%) |
| **Level of support provided to manage personal hygiene during periods at home** |  |
| Completely independent | 2 (2.9%) |
| Indirect support | 20 (28.6%) |
| Partially assisted | 27 (38.6%) |
| Fully assisted | 21 (30.0%) |
| **Level of support provided to manage personal hygiene during periods at school/college** |  |
| Completely independent | 16 (22.9%) |
| Indirect support | 28 (40.0%) |
| Partially assisted | 15 (21.4%) |
| Fully assisted | 11 (15.7%) |
| **Frequency of leakage of menstrual blood** |  |
| Never | 19 (27.1%) |
| Some of the time | 45 (64.3%) |
| Most of the time | 4 (5.7%) |
| All of the time | 2 (2.9%) |

**Supplemental Table 5. Areas of demand for menstruation support**

| **Information sources and topics that would be valued going forwards** | **Young person not yet menstruating**  **(N=23)** | **Young person is menstruating**  **(N=120)** |
| --- | --- | --- |
| Finding resources specifically designed for / accessible to young people with learning disabilities | 18 (78.3%) | 53 (44.2%) |
| Strategies to support your child/young person’s (independent) management of their periods | 19 (82.6%) | 45 (37.5%) |
| Materials / resources to share with your child/young person about periods | 18 (78.3%) | 39 (32.5%) |
| Materials / resources to share with your child/young person about the menstrual cycle | 12 (52.2%) | 31 (25.8%) |
| Information on managing menstrual pain | 7 (3.0%) | 35 (29.2%) |
| Information on different menstrual products | 8 (3.5%) | 20 (16.7%) |
| Information about hormonal changes throughout the menstrual cycle | 7 (3.0%) | 29 (23.2%) |
| Information on hormonal medication | 5 (2.2%) | 31 (25.8%) |
| Information on contraceptives | 4 (1.7%) | 19 (15.8%) |
| Information relevant to specific cultural or religious aspects of menstruation | 2 (0.9%) | 8 (6.7%) |
| None of the above - I do not need any information or support | 1 (0.4%) | 29 (24.2%) |
